# Anti-racism in postgraduate medical curricula

**DOI:** 10.1101/2025.01.22.25320921

**Authors:** Chetna Sharma, Hadjer Nacer, Aaron Koay, Martin McKee, Delan Devakumar

## Abstract

Racism is a public health threat and is firmly ingrained in the healthcare system. The reasons for this are complex as racism is rooted in historical, political, economic and social structures of society. To improve the health of their patients and the public, doctors must be able to recognise, name and act on the impact of racism. This study aims to understand action taken by postgraduate medical organisations in the UK in the aftermath of the global public reckoning of structural racism and its longstanding impacts on societal health in 2020. We analysed the public statements made by postgraduate medical organisations and then how this relates to the learning outcomes that doctors in training need to complete. We found that while many institutions (n=14) issued public commitments to anti-racism, reviewing the content of all GMC-approved postgraduate curricula (n=102) shows that the majority of UK doctors are receiving minimal or no education or training in anti-racism. As such, we call institutions involved in postgraduate medical education to include anti-racism competencies in medical curricula to support doctors to develop the skills, behaviours and knowledge to name and address the role of racism in health inequalities.

## Introduction

Racism is established as a determinant of physical and mental health, with significant associations that persist after adjusting for other characteristics.^1^ The pathways through which they act are increasingly understood,^2,3^ with structural discrimination seeping into the functioning of institutions and systems, including health systems, subsequently impacting on health.^4^ Structurally embedded discrimination perpetuates disparities in power, constraining access to resources and opportunities, which, in turn, cause health inequalities. In the UK, adults from minoritised ethnicities who have faced racial discrimination are more likely to experience limiting longstanding illness, poorer physical and mental functioning, greater psychological distress, worse life satisfaction and poor self-rated health, after adjusting for age, sex, and socioeconomic factors including household income and education.^5^ The recent national review of child mortality shows one of the starkest examples of racial health inequalities. Reviewing data over the last five years, infants of Black Caribbean or Black African ethnicity are over three times more likely to die than infants of White ethnicity.^6^ A report revealed that the average age of death for people with a learning disability who are from an ethnic minority background is 34 years, just over half the life expectancy of their White counterparts at 62 years of age, with the report highlighting potential factors influencing this, including poorer healthcare access, experience and outcomes.^7^. Racial bias, however, is not always about measurable health outcomes; it can be as simple as the language used to describe patients. A study that reviewed health records found that Black patients are more than twice as likely to receive negative descriptors, such as ‘resistant’ or ‘non-compliant’ compared to White patients.^8^

Although these health inequalities have long been felt and known, they were catapulted to mass public consciousness by the COVID-19 pandemic in 2020, with its disproportionate impact on racially minoritised populations, coinciding with the increased awareness of the Black Lives Matter Movement following the murder of George Floyd in May of that year. This shone a light on structural racism in many UK institutions,^9^ with recognition that centuries of racial discrimination is hard-wired in the NHS. The Workforce Race Equality Standard (WRES)^10^ and, more recently, the Medical Workforce Race Equality Standard (MWRES)^11^ revealed disparities in career progression, experience, and opportunities that exist for people from minoritised populations compared with White staff. The NHS Race and Health Observatory was also set up in 2021 to examine and tackle inequalities experienced by Black and ethnically minoritised patients, communities and staff.^12^

There has concurrently been a push toward improving equality, diversity and inclusion (EDI) in organisations. While EDI aims to bring about equality within the workforce and treatment of individuals, more intentional education and action is needed to understand the underlying causes of these inequalities and reduce patient and population inequities. There is an important distinction between anti-racism work, and the EDI agenda. Anti-racism can be defined as ‘the active process of identifying and eliminating racism by changing systems, organizational structures, policies and practices and attitudes, so that power is redistributed and shared equitably’.^13^ Anti-racism provides the foundational understanding that it is not race, ethnicity, religion, or migratory status that deems an individual to become unwell or fare differently. Rather it is the way the society in which they live and work, places value on them because of their characteristics, with the associated unequal power dynamics influencing their health outcomes and access to resources and opportunities. As healthcare professionals, we must acknowledge the racist and discriminatory ideologies and practices that underlie modern medicine and ensure that future practice views the structures that underpin health systems through a critical lens, to enable us to provide the best quality care for all patients.

This calls for the incorporation of specific learning outcomes within curricula that go beyond generic acknowledgement of the problem, to educate and equip doctors with the knowledge and skills for applying antiracist approaches in their practice.^14^ This must acknowledge that doctors, like anyone else, may already hold misconceptions about race and ethnicity^15–18^ that are likely to perpetuate disparities.^19,20^ In the context of widening health inequalities in the UK, there is a clear need for medical education to include knowledge of discrimination as a determinant of health^21^, conferring the sensitivity and skills to identify structural racism^22,23^ and to address it.^24–26^

This raises the question of the extent to which this is already happening. To answer it, we undertook two analyses. Firstly, we reviewed publicly available commitments from Royal Colleges, Faculties and examination bodies to address racism, xenophobia, religious discrimination (explicitly including Islamophobia and antisemitism given contemporary discourse)^27–29^ and decolonisation, and whether the commitments included education-related actions. Decolonisation was added as a concept given the rise in its use as an approach to address inequalities resulting from colonial injustices, therefore including it in the analyses allowed us to investigate the type of actions outlined to address these structural harms. We also sought evidence of any stated measures of accountability.^30^ Secondly, we reviewed postgraduate medical curricula to identify whether the impact of racism and discrimination on health was explicitly mentioned as one of the competencies expected of doctors in training.

## Methods

We limited our analyses to medical professionals in the UK (England, Northern Ireland, Scotland, and Wales). In the UK, postgraduate education and training of health professionals is provided by approved Faculties and Royal Colleges using curricula that they develop for their respective specialties.^31^ The General Medical Council (GMC), the medical regulator, approves these curricula against standards set out in ‘Excellence by design: standards for postgraduate curricula’ and the supplementary ‘Generic professional capabilities framework’.^32,33^ In some cases, these processes are combined across multiple specialties; for example the Joint Royal Colleges of Physicians Board is responsible for training and curriculum design for 31 specialties. The list of organisations responsible for the 102 curricula is in Appendix 1.

The analysis was based on materials that were publicly available up to August 2023. The search terms used were *race, racism, ethnic, ethnicity, xenophobia, religion, religious, Islamophobia, antisemitism* and *decolonising.* We analysed the websites of 21 organisations identified as being active in postgraduate medical education, to review their public commitments. Searches were conducted using the Google ‘site:’ command and website-specific search engines. Statements were included if they were explicit commitments, policies, action plans or strategies to address racism, xenophobia, Islamophobia, antisemitism or decolonisation. Guest blogs, webinars, events and research papers were excluded. Generic equality, diversity and inclusion statements (e.g. stating that the Equality Act will be upheld) were excluded, as these are minimum legal requirements. Particular attention was paid to commitments that considered intersectionality, i.e., interactions of race or ethnicity with other social positions such as gender or disability. Intersectionality is the understanding that discrimination based on differing forms of societal hierarchy and oppression (e.g., race, religion, gender or class) are not neatly additive, but instead lead to a reconstituted impact, which can manifest in health inequalities. Measures involving education and training within these public commitments were charted, as was any evidence of accountability mechanisms.^30^

In the review of curricula, we additionally analysed for mention of *determinants* to identify learning outcomes that acknowledged structural and wider conditions impacting health. The GMC-approved postgraduate curricula list was used as the sampling frame for curricula. All 65 GMC-approved speciality curricula, 6 core curricula and 31 sub-speciality curricula (total n=102 postgraduate curricula) were reviewed. Searches were carried out on the GMC website search engine. Terms used were differentiated on whether they related explicitly to a learning outcome or competency or whether they were used generically, for example in a section on equality & diversity without a direct link to an educational learning point.

Items that included individual search terms were extracted verbatim into Excel spreadsheets, with double coding by two researchers (CS and HN). Discrepancies were resolved by discussion or by a third researcher (AK). Of those that included mention of race or racism specifically within learning outcomes, inductive analysis was carried out to identify themes. The results remain the interpretations of available information as understood by the authors. To ensure any relevant material had not been missed, educational leads at the Royal Colleges and Faculties were emailed the data that pertained to their speciality to confirm that the information found publicly was correct.

The UCL Research Ethics Committee was consulted during project conception. It advised that ethics approval was not required as only publicly available information was used and individuals are not named.

## Results

Results are described in two parts. Firstly, we outline the findings of our analysis of public statements and commitments, for all 21 organisations identified as responsible for postgraduate medical education in the UK. An example is given of a public commitment, which has an education-related measure, along with an accountability mechanism for this measure. Secondly, we present the analysis of all GMC-approved postgraduate curricula, specifically identifying relevant terms within learning outcomes. An inductive analysis of curricula that mention race or racism as part of a learning competency reveals themes across specialities.

### Statements and commitments

All 21 organisations responsible for postgraduate medical curricula were investigated for public commitments to anti-racism, and addressing xenophobia, religious discrimination (including Islamophobia and antisemitism), and decolonisation. Figure 1 displays identified relevant public commitments, any education-related measures for those commitments, followed by any accountability mechanisms to ensure these measures are actioned. 16 organisations (76%) had at least one relevant public commitment, of which 14 commitments (88%) related to anti-racism and 4 (25%) related to religion. Three organisations (14%) - the Faculty of Public Health, the Royal College of Obstetricians and Gynaecologists, and the Royal College of Psychiatrists - also mentioned intersectionality.

**Figure 1.**
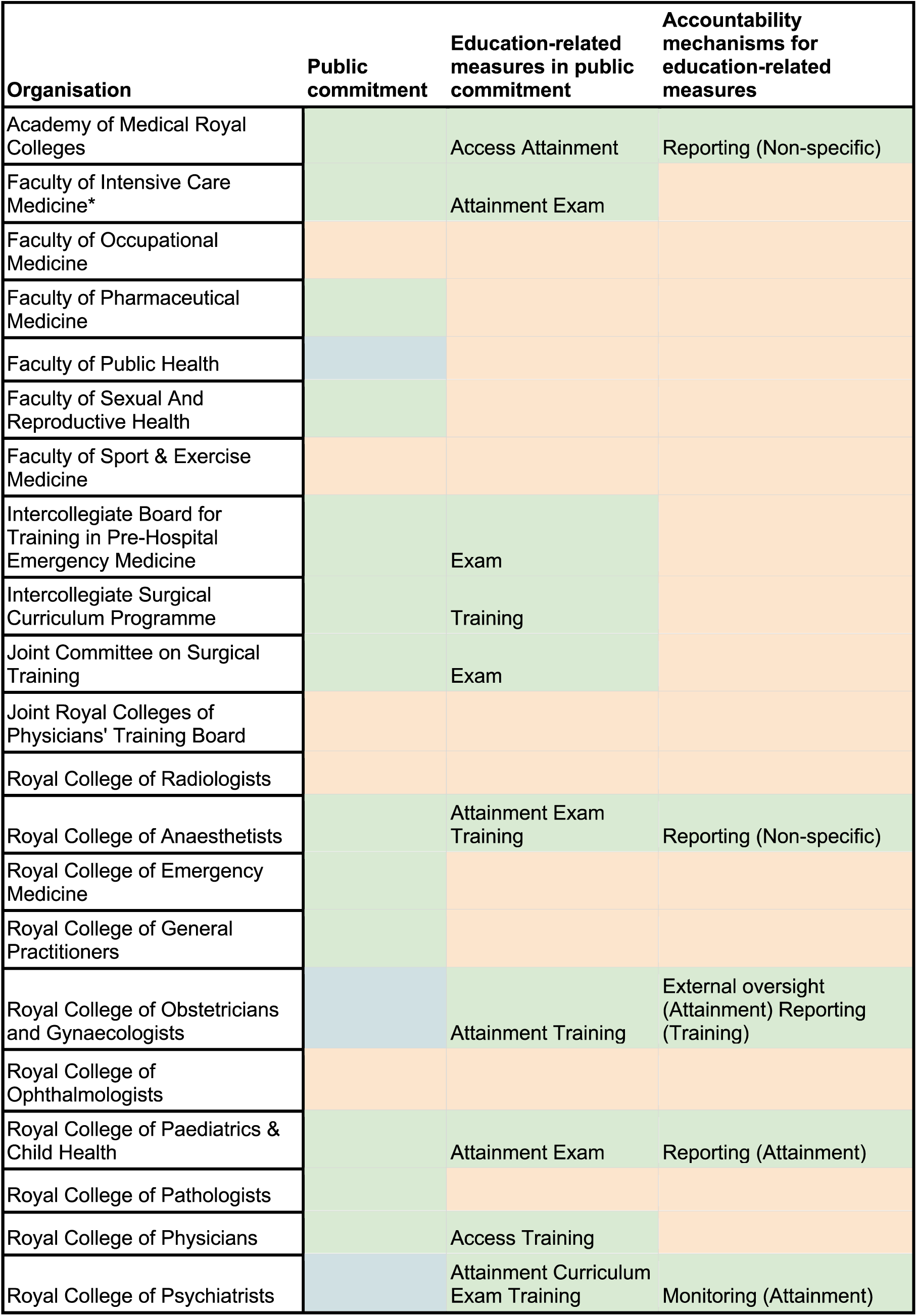
Public commitments by Royal Colleges, Faculties and other training bodies. Green: relevant public statement identified; Orange: not found; Blue: takes an intersectional lens; *: organisations that responded to data verification for public statements. Text in boxes refer to the type of education-related measure and type of accountability measure that was identified.

Ten organisations (48%) mentioned education-related measures within their public commitments, of which only the Royal College of Psychiatrists mentioned curriculum content. Other education-related measures pertained to the workforce, focusing on attainment, access, exams and training. Five organisations (24%) included education-related measures linked to an accountability mechanism, most pertaining to reporting. Panel 1 provides an example from the Royal College of Psychiatrists.

##### Panel 1. An example of a public commitment – Royal College of Psychiatrists

###### Public commitment

*“In order to put together an ambitious new race equality strategy, which will look closely at the training curricula, practice guidelines and much else, I [President] am putting together a Race Equality Taskforce.”*^34^

###### Education-related measures in public commitment

Curriculum: *“Reviewing the core and higher training curricula to ensure they adequately reflect the knowledge and skills required to deliver clinical care that is equitable for all, including understanding the impact of structural inequalities and power differentials within mental health”*

Attainment: “*Pilot and evaluate training, support and engagement activities to inform further initiatives to tackle differential attainment”*^34^

###### Accountability mechanisms for education-related measures

*Monitoring (Attainment):* “*The College monitors differential attainment after each MRCPsych examination”*^34^

### Curricula and learning outcomes

All 102 postgraduate curricula had been updated in 2021 or later. (Organisations with corresponding speciality curricula can be found in Appendix 1). The percentage of curricula mentioning each of the key terms is shown in Figure 2. We differentiated between the mention of a term within a learning competency, which trainees are assessed on, compared to their generic mention within the curricula text. Eleven curricula (11%) mentioned “racism”, including ten as part of a learning outcome. However, eight of these were for sub-specialities covered by the Royal College of Psychiatrists; the other two were from the Faculty of Public Health and the Royal College of Paediatrics and Child Health. Therefore, only three specialities included racism and its impacts on health within a learning outcome. Sixteen curricula (16%) mentioned “race” (rather than “racism”), but only three included it within a learning outcome or competency. Twenty-six (25%) used the terms ethnic or ethnicity, with thirteen including it within a learning outcome. All mentioned collecting data on ethnicity of trainees and taking account of ethnicity when interacting with patients, while seven (7%) discussed the role of ethnicity in the clinical presentation. Twenty-two (22%) mentioned religion, with nine including it in a learning outcome. Twenty-two (22%) mentioned broader health determinants and five (5%) mentioned intersectionality (of which three were subspecialties of the Royal College of Psychiatrists and the others the Royal College of Paediatrics and Broad Base Training by AoMRC), all were included in learning outcomes. No curriculum contained any mention of xenophobia, antisemitism, Islamophobia or decolonisation.

**Figure 2.**
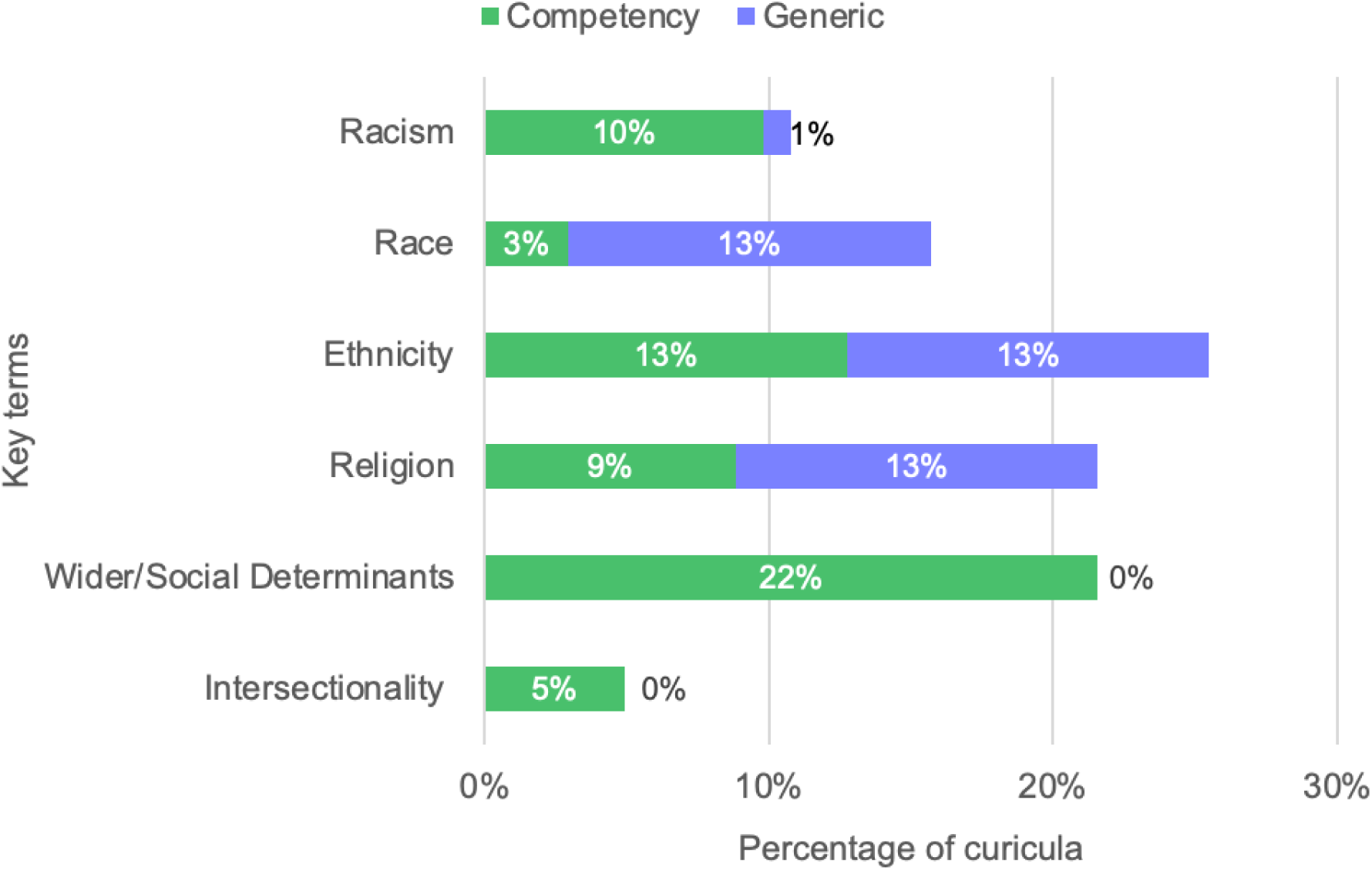
Percentage of curricula mentioning key terms, differentiated by their inclusion within a learning competency or generic mention in the curricula text.

Five specialities replied to our requests to check data, no inaccuracies were reported. Responses included the following additional material: information on a decolonising workstream as part of a curriculum update initiative; development of professional guides that cover wider determinants of health inequalities to support the core curriculum; information on an EDI assessment of the impact of the curriculum; and on generic EDI-related education. Most curricula mentioned equity, diversity, and inclusion in the generic text to signify the college or faculty’s efforts to advance EDI principles.

An inductive analysis of the learning outcomes that explicitly mentioned race and racism showed considerable variation in how organisations used the terms (Table 1). The Faculty of Public Health defined racism as a public health issue and included it as an overarching theme of the curriculum, with trainees being expected to develop an understanding and consider approaches to mitigate widening health inequalities. The Faculty of Public Health, the Royal College of Psychiatrists and Paediatrics included an understanding of the impacts of racism on health as a learning competency for trainees. The final theme we identified mentioned differences in disease burden related to race, however without context for underlying or structural factors producing these differences.

**Table 1.** Representation of race and racism in Learning Outcomes and High-Level Outcomes.

| Theme | Learning Outcomes/ High Level Outcomes | Examples of how a learning outcome may be met |
| --- | --- | --- |
| Naming and defining racism as a social determinant of health | **Faculty of Public Health:** “Racism is a public health issue, and it is vital that as part of the curriculum and their ongoing learning, registrars consider approaches to mitigate any further widening of inequalities amongst ethnic minority communities.” | No examples |
| Including anti-racism understanding as a competency and as a learning outcome for registrars | **The Royal College of Psychiatrists:** “Demonstrate an understanding of individual variation and the impact of social, cultural, spiritual and religious factors, including effects of deprivation, discrimination and racism.”<br>**Faculty of Public Health:** “Demonstrate cultural competence and is able to work effectively in cross- cultural situations both internally and externally to the organisation: The registrar will demonstrate an understanding of how structural determinants to health, such as systemic racism, influence the development of culturally competent policies.” | **Faculty of Public Health:** “The registrar to produce position statement on structural racism as a determinant of health.”<br>**Paediatrics:** “15-year trans old boy is seen in ED. The trainee uses their understanding of both racism and transphobia as social determinants of health to form an intersectional understanding of their physical and mental health.” |
| Presenting disease burden based on race without context | **Endocrinology and Diabetes** by JRCPTB: “Understanding of racial variations in diabetes and endocrine disease including presentation, complication and management” |  |

## Discussion

This study is the first to consider how anti-racism is represented in postgraduate medical curricula in the UK. We reviewed public statements related to anti-racism as these set out the course and vision of an organisation and should drive action. We then analysed postgraduate curricula to determine how an understanding of structural discrimination has translated into medical education and competencies expected of doctors.

Most organisations responsible for postgraduate medical curricula made some public commitment to anti-racism, a welcome initial sign of commitment to act. Over a third included education-related measures, all relating to attainment, access, exams, and training, something that is also to be welcomed given patterns of disparities in educational attainment amongst postgraduate doctors in the UK. A recent GMC report showed continued differential attainment by ethnicity, with inequalities in exam pass rates, reviews of competence and career progression among doctors of racially minoritised backgrounds compared to those of White backgrounds. These disparities were compounded by other factors including socioeconomic background, faith and disability.^35^

Yet, even with two-thirds of the organisations making some public commitment related to anti-racism, of which over half included education-related measures, our analysis of all 102 curricula, showed only ten (from three distinct specialities) included the impact of racism on health within the competencies expected of their trainees. None named health outcomes in relation to xenophobia, antisemitism, Islamophobia or commented on decolonisation, a major gap. Of the curricula that mentioned race, the majority were in the context of fair treatment of individuals based on protected characteristics within Equality and Diversity legislation, but few extended to an understanding of the underlying processes leading to unequal health outcomes.

We reviewed the global literature on anti-racism in postgraduate medical education (Panel 2). The Royal College of Physicians and Surgeons of Canada, currently updating their 2025 Physician Competency Framework,^36^ provides an example of good practice. Members of the steering committee have expressed that ‘all physicians in Canada must demonstrate ongoing competence in anti-racist, anti-oppressive praxis, to address the impact of racism both on patient outcomes and on their physician colleagues who identify as Black, Indigenous, or people of color’.^37^ As part of the competency framework update, an extensive literature review was conducted that proposed ten themes that should be incorporated into postgraduate medical competencies, including anti-racism, equity, diversity, inclusion, and social justice.^36^ The processes of educational change and subsequent implementation must be inclusive and collaborative, engaging with all who have a stake, so that work under the banner of anti-racism does not reproduce the power dynamics that contributed to health inequalities in the first place. Prince and colleagues, reflecting on medical training, call for measures to imbue the next generation of doctors with a desire for lifelong learning and advocacy based on a three-pillar framework that includes trust building, structural competency, and cultural humility.^22^

##### Panel 2 Research in context

###### Literature search

A literature search was conducted to identify articles related to anti-racism in postgraduate medical education. (See Appendix 2 for PRISMA flowchart). We searched PubMed and Google Scholar using the search strategy:

(((racism OR antiracism) AND (training OR education OR competencies OR competency OR syllabus OR syllabi OR curricula OR curriculum) AND (postgraduate OR doctor OR clinician OR physician) NOT (undergraduate OR medical student))).

From 709 references yielded by our search, 23 publications were eligible for inclusion. Most of the publications were after 2020 (n=20), with fifteen publications from the USA, four from Canada and one from Australia. All competencies had a target audience of training clinicians/residents with 14 articles focused on all specialties, 9 aimed at specific specialties (Internal Medicine n=1^38^, Emergency Medicine n=1^25^, Infectious Diseases n=1^24^, Psychiatry n=2^39,40^, Paediatrics n=3^14,41,42^, Family Medicine n=1 ^43^), which were still included due to their applicable competencies. Overall, all articles referred to the need for inclusion of anti-racism competencies in postgraduate medical education. Eleven articles referred to the need to name, recognise and acknowledge racism as a determinant of health. Seven articles proposed and evaluated approaches to anti-racism education, for example through strategies to improve physician-patient communication,^44^ developing an antibias curriculum, ^45^ using community organising and public narrative approaches, ^46^ and frameworks for anti-racism education. ^24,36^

###### Interpretation

Overall, findings highlight the need to include anti-racism education in postgraduate medical education, starting with naming, recognising and acknowledging racism as a determinant of health. The literature search provides examples of approaches and methodologies for anti-racism education in postgraduate medical education but highlights a lack of comprehensive frameworks for core anti-racism competencies and learning outcomes.

Given our findings, what must be done to accelerate meaningful anti-racism reforms in postgraduate medical education? This paper has examined the situation in one country, the UK. However, it is likely that the gaps we identified are present in many other countries and our first recommendation is that our study should be replicated elsewhere.

Our remaining recommendations are directed at the key actors in the UK, but we hope that once similar exercises have been conducted elsewhere, they could be adapted to other contexts. For this to happen it will be necessary to map the roles of the various actors involved. Even within Europe the regulation, governance, and representation vary greatly.^47,48^ For example, the British Medical Association is both a trade union and a professional association while these roles are separate in some other countries. Entities with similar profiles to the British Royal Colleges can be found in Ireland and in some Commonwealth countries but are rare elsewhere. The role of regulators varies greatly.^49^ Hence, in directing our recommendations to particular bodies, we note that we are referring to the roles that they fulfil in the UK and readers in other countries will have to map these roles onto their own structures. Noting this important constraint, we offer the following recommendations.

### 1 Royal Colleges and Faculties responsible for postgraduate curricula

Royal Colleges and Faculties need to rethink, reframe, and reconstruct their curricula with an anti-racist lens. Naming racism as a problem is a first step but much more must be done. Measures include achieving precision in terminology, for example avoiding conflation of race and ancestry by means of imprecise labels. Training in structural racism equips doctors to understand how social, political, and historical forces have influenced and impacted health. Anti-racist education must make systemic oppression visible, recognise personal complicity and develop strategies that can address structural inequalities. It is also crucial to contextualise differences in racial and ethnic disease burden within wider structural and social determinants of disease.

### 2. GMC as the medical regulator responsible for setting standards and approving curricula

The GMC needs to ensure that anti-racism is an integral part of postgraduate training programmes. This aligns with the inclusion of tackling discrimination based on personal characteristics, including explicit mention of race and religion, in the 2024 update of Good Medical Practice, which sets out the expectations of all doctors registered with it. The next curriculum update, in 2025, provides an opportunity to do this.

### 3. Academy of Medical Royal Colleges and British Medical Association, as bodies that can address issues where a cross-specialty perspective is needed, ideally working together to do so

The Academy provides a forum for the Medical Royal Colleges to support educators with practice-based, peer-reviewed resources and pedagogies for anti-racist knowledge and curricula. The Association of American Medical Colleges, the equivalent of AoMRC in the United States, has developed a collection of anti-racism education resources to support colleges with practice-based, peer-reviewed resources to teach anti-racist knowledge and clinical skills. It also promotes scholarship on anti-racist curricula convenes a community of collaborators dedicated to the elimination of racism within medical education.^50^ The British Medical Association, with its long history of leadership on medical ethics, can develop and promote standards for doctors in their everyday practice.

### Limitations

This study is the first to explore anti-racism in UK postgraduate medical curricula. However, our findings only apply to the curricula we analysed. We cannot exclude the possibility that there may be other local guidance issued by other bodies, recognising that specialist postgraduate medical training routes are complex and influenced by many organisations and stakeholders including NHS England and Local Training Boards. It is also a snapshot in time, and we note that some specialties are developing resources or have set up taskforces that could change the situation in the future. We relied on what is written, and it is possible that anti-racism is included informally in some training programmes. For example, many public health trainees undertake a MSc as part of their training, which can incorporate an understanding of structural determinants of health.

We assessed discrimination based on race, ethnicity, religion and migratory status and we acknowledge there are multiple other forms of discrimination that often intersect. These social identities were selected to be in line with those investigated in the Lancet Series. We were limited to information that is available on organisational websites and so further details that exist not in the public realm will not be accounted for.

## Conclusion

A multifaceted approach is needed to address the health harms of racism and structural discrimination that manifest at all levels from the individual to the societal and over time, recognising that doctors play a critical role in tackling these inequalities. Through a review of publicly available information, we have been able to understand and analyse public-facing messaging from medical speciality associations in the UK and the postgraduate competencies expected in these specialities. We identified a lack of comprehensive anti-racism education, with a long way to go to capture the impact of discrimination based on race, ethnicity, religion, and migration on health. There is a legal and policy imperative for doctors to have the corresponding competencies, as well as a stated desire by Royal Colleges to understand how to do this more effectively. There is a need for cross-organisation learning and collaboration to develop an anti-racist framework that defines the skills and level required at different points in training. Improved evaluation of programme delivery would help identify successful practices to share within and across Royal Colleges and Faculties, while maintaining flexibility of provision. Co-creation with patients and the public is vital to ensure integrity in this approach, redress unequal power dynamics and tackle health inequalities. Importantly, this includes collaborative work around how curricula are interpreted in practice, across patients, trainees and trainers. This points to the necessity for future work in understanding how educational change is enacted, with measures of success in antiracist education to be determined in collaboration with communities who are impacted.

## Supporting information

Appendix 1: Supplemental Table 1. Organisations and corresponding medical postgraduate speciality curricula

Appendix 2: Supplemental Figure 1. PRISMA flow diagram of search process

## Data Availability

All data produced in the present study are available upon reasonable request to the authors

