## Appendix 1: Supplemental Table 1. Organisations and corresponding medical postgraduate speciality curricula for "Anti-racism in postgraduate medical curricula"

| **Organisations responsible for training and setting curricula** | **Training/specialty** |
|---|---|
| Academy of Medical Royal Colleges | Broad based training |
| Faculty of Intensive Care Medicine | Intensive care medicine |
| Faculty of Occupational Medicine | Occupational medicine |
| Faculty of Pharmaceutical Medicine | Pharmaceutical medicine |
| Faculty of Public Health | Public health medicine |
| Faculty of Sexual And Reproductive Health | Community sexual and reproductive health |
| Intercollegiate Board for Training in Pre-Hospital Emergency Medicine | Pre-hospital emergency medicine |
| Joint Committee on Surgical Training (parent organisation for Intercollegiate Surgical Curriculum Programme) | Cardio-thoracic surgery<br>General surgery<br>Neurosurgery<br>Oral and maxillo-facial surgery<br>Otolaryngology<br>Paediatric surgery<br>Plastic surgery<br>Trauma and orthopaedic surgery<br>Urology<br>Vascular surgery<br>Congenital cardiac surgery<br>Core surgical training |

| | |
|---|---|
| Joint Royal Colleges of Physicians' Training Board (includes Faculty of Sport & Exercise Medicine and Royal College of Physicians) | Acute internal medicine<br>Allergy<br>Audio vestibular medicine<br>Aviation and space medicine<br>Cardiology<br>Clinical genetics<br>Clinical neurophysiology<br>Clinical pharmacology and therapeutics<br>Dermatology<br>Endocrinology and diabetes mellitus<br>Gastro-enterology<br>General (internal) medicine<br>Genito-urinary medicine<br>Geriatric medicine<br>Haematology<br>Immunology<br>Infectious diseases<br>Medical oncology<br>Medical ophthalmology<br>Metabolic medicine<br>Neurology<br>Nuclear medicine<br>Paediatric cardiology<br>Palliative medicine<br>Rehabilitation medicine<br>Renal medicine<br>Respiratory medicine<br>Rheumatology<br>Sport and exercise medicine<br>Stroke medicine<br>Tropical medicine |
| Royal College of Radiologists | Clinical oncology<br>Clinical radiology<br>Interventional radiology |
| Royal College of Anaesthetists | Anaesthetics<br>Core anaesthetics |
| Royal College of Emergency Medicine | Emergency medicine |
| Royal College of General Practitioners | General practice |
| Royal College of Obstetricians and Gynaecologists | Gynaecological oncology<br>Maternal and fetal medicine<br>Obstetrics and gynaecology<br>Reproductive medicine<br>Urogynaecology |
| Royal College of Ophthalmologists | Ophthalmology (Not available online) |

| | |
|---|---|
| Royal College of Paediatrics & Child Health | Child mental health<br>Community child health<br>Neonatal medicine<br>Paediatric allergy, immunology and infectious diseases<br>Paediatric clinical pharmacology and therapeutics<br>Paediatric diabetes and endocrinology<br>Paediatric emergency medicine<br>Paediatric gastroenterology, hepatology and nutrition<br>Paediatric inherited metabolic medicine<br>Paediatric intensive care medicine<br>Paediatric nephrology<br>Paediatric neurodisability<br>Paediatric neurology<br>Paediatric oncology<br>Paediatric palliative medicine<br>Paediatric respiratory medicine<br>Paediatric rheumatology<br>Paediatrics |
| Royal College of Pathologists | Chemical pathology<br>Cytopathology<br>Diagnostic neuropathology<br>Forensic histopathology<br>Histopathology<br>Medical microbiology<br>Medical virology<br>Paediatric and perinatal pathology |
| Royal College of Psychiatrists | Addiction psychiatry<br>Child and adolescent psychiatry<br>Core psychiatry training<br>Forensic psychiatry<br>General psychiatry<br>Liaison psychiatry<br>Medical psychotherapy<br>Old age psychiatry<br>Psychiatry of learning disability<br>Rehabilitation psychiatry |
