## Appendix 2: Supplemental Figure 1. PRISMA flow diagram of search process for "Anti-racism in postgraduate medical curricula"

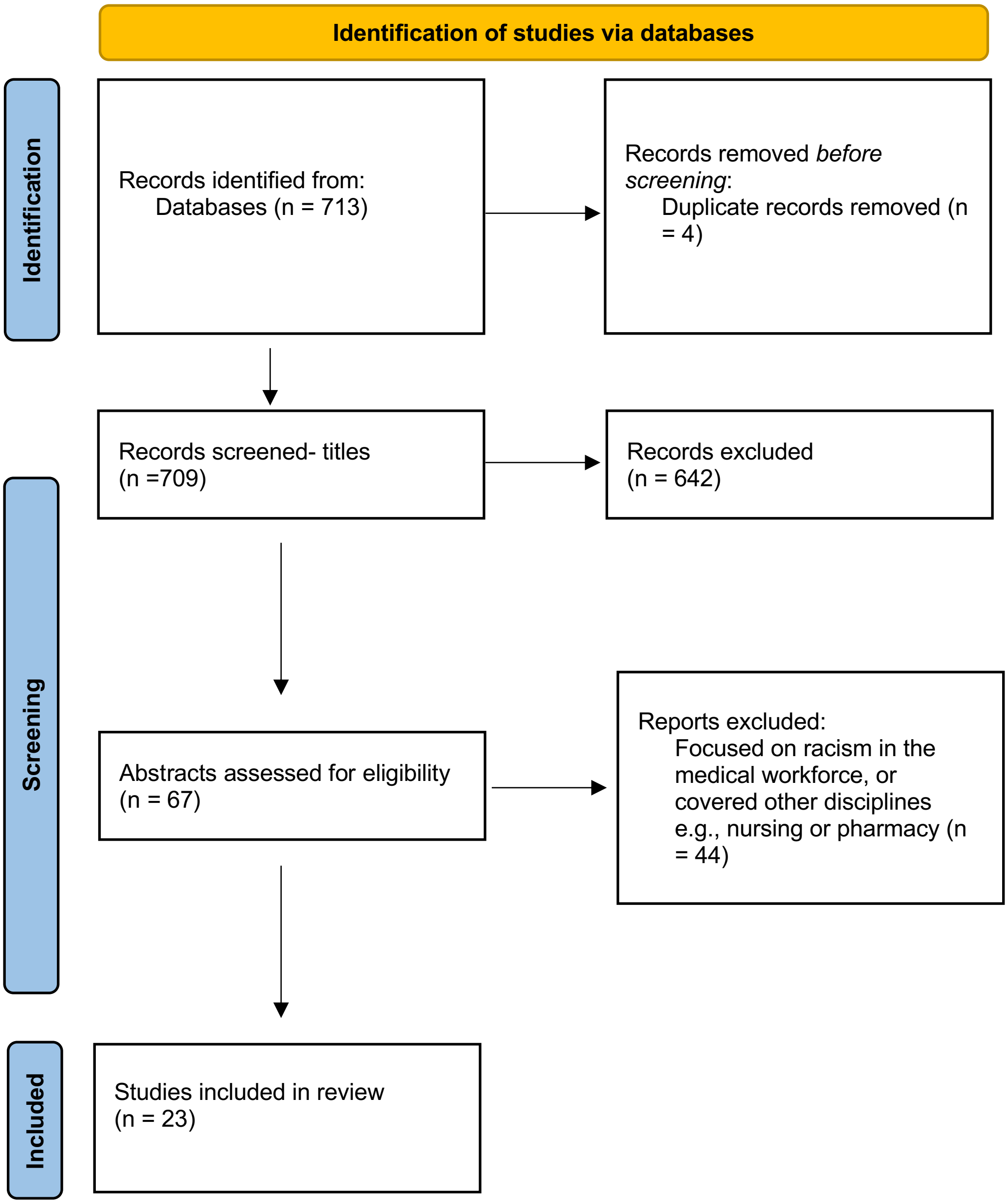
